# Cognitive and brain reserve in bilingual speakers with clinical AD variants

**DOI:** 10.64898/2026.03.27.26349575

**Authors:** Nicoletta Biondo, John Matthew Suntay, Muskaan Sandhu, Jeanine Sandra Estaban, Janhavi Pillai, Maria Luisa Mandelli, Eugenie Mamuyac, Rochelle-Jan Reyes, Ariel Guevarra, Maya L. Henry, Nina F. Dronkers, Maria Luisa Gorno Tempini, Stephanie M. Grasso, Jessica de Leon

**Author notes:** Corresponding author: Jessica de Leon, MD, Memory and Aging Center, Department of Neurology, University of California, San Francisco (UCSF). Stephanie M. Grasso and Jessica de Leon contributed equally to this study.

## Abstract

**INTRODUCTION:** Bilingualism may confer resilience via enhanced neural integrity. However, evidence for bilingualism’s neuroprotective effect is mixed, and studies across Alzheimer’s disease (AD) variants are scarce. This study examined gray matter volume (GMV) differences between bilinguals and monolinguals with amnestic AD and logopenic variant primary progressive aphasia (lvPPA).

**METHODS:** In 136 amnestic AD and 88 lvPPA participants with neuropsychological assessments and structural MRI, we analyzed differences between monolinguals and bilinguals within each variant, controlling for demographic covariates.

**RESULTS:** Amnestic AD bilinguals exhibited less GMV in hippocampal, fusiform, and occipital regions compared to monolinguals. LvPPA bilinguals had less temporal and occipital volumes, but they had greater volumes in inferior parietal regions, which are considered a disease epicenter in lvPPA. Cognitive performance in monolinguals and bilinguals was comparable within variants.

**DISCUSSION:** Bilingualism may support cognitive reserve (preserved cognition despite reduced GMV) in both AD variants, with additional brain reserve in lvPPA.

## BACKGROUND

The escalating global prevalence of neurodegenerative diseases, particularly Alzheimer’s disease and related dementias (ADRD), poses a formidable challenge to public health and societal well-being. Characterized by progressive cognitive decline and significant neuropathological changes, these conditions underscore an urgent need to identify modifiable factors that might confer resilience or delay symptom onset. Among the various lifestyle and environmental influences investigated, bilingualism has emerged as a compelling area of research, with a growing body of evidence suggesting its potential role in shaping cognitive trajectories and brain integrity across the lifespan, especially in the context of aging and neurodegeneration.

The concept of cognitive reserve posits that individuals with cognitively enriching life experiences, such as education, engaging occupations, or complex leisure activities, can better withstand brain pathology before manifesting clinical symptoms of dementia.[1] This framework suggests that a more efficient or flexible cognitive network can compensate for underlying neuropathological damage, allowing individuals to maintain cognitive function (‘resilience’) despite accumulating brain changes.[2–4] Closely related, the concept of brain reserve refers to the structural or quantitative aspects of the brain, such as larger brain volume, greater neuronal count, or synaptic density, which may provide a buffer (‘resistance’) against the effects of pathology.[2–6] In essence, brain reserve represents the brain’s physical capacity or “hardware,” the sheer amount of neural capital available, while cognitive reserve reflects the efficiency and adaptability of its “software” or cognitive processes, enabling the brain to utilize its resources more effectively or recruit alternative networks when faced with damage. Higher educational attainment, in particular, has consistently been associated with delayed onset of dementia symptoms, greater efficiency in neural network recruitment, and preserved cognitive performance despite comparable levels of neuropathology.[1,7] Similarly, bilingualism, defined as the regular use of two or more languages, is hypothesized to contribute to both cognitive and brain reserve by continuously engaging executive functions, such as attention, inhibition, and task-switching, which are crucial for managing multiple language systems.[8] This lifelong cognitive training is thought to foster neural adaptations that enhance brain reserve and maintenance, potentially delaying the clinical expression of neurodegenerative diseases.

Studies in healthy bilingual adults have revealed distinct structural differences in brain morphology compared to monolinguals, reflecting the brain’s remarkable neuroplasticity in response to sustained linguistic demands. Early pioneering work, such as that by Mechelli et al., (2004)[9] demonstrated greater gray matter density in the left inferior parietal cortex of bilingual compared to monolingual individuals, a region critically involved in language processing[10] and second language learning.[11] Volume in this region was more pronounced in individuals who acquired their second language earlier and correlated positively with proficiency, underscoring the experience-dependent nature of these changes. Subsequent research has expanded on these findings, showing that bilingualism is associated with differences in gray matter volume (GMV) and cortical thickness (CT) in a distributed network of brain regions. These include regions, often but not always bilaterally, within the frontal lobe, particularly the dorsolateral prefrontal cortex, implicated in executive functions; the anterior cingulate cortex, involved in conflict monitoring; and various temporal and parietal regions, which are central to language and semantic processing.[12,13] The extent and localization of these structural differences often correlate with factors such as age of second language acquisition, proficiency, and the intensity of language use, suggesting a dynamic interplay between linguistic experience and brain plasticity.[14] While some studies report greater GMV or CT in bilinguals, indicative of greater neural resources, others note smaller or thinner brain regions despite similar performance compared to monolinguals, thus suggesting that bilingual brains may have a more efficient and streamlined neural architecture.[14,15] These structural adaptations are thought to underpin the observed cognitive differences observed in bilinguals, particularly in tasks requiring executive control.

A growing body of research suggests that lifelong bilingualism may delay the onset of dementia symptoms, including those of Alzheimer’s dementia (AD), by several years, typically ranging from 4 to 5 years.[16,17] This delay is often observed despite equal or greater levels of neuropathology, such as greater brain atrophy (i.e., reduction in brain tissue due to neuronal loss and synaptic degeneration) or hypometabolism, in the brains of bilingual relative to monolingual individuals at the time of clinical diagnosis, lending support to the cognitive reserve hypothesis. This finding suggests that bilingualism enhances the brain’s ability to tolerate a greater amount of neuropathology before clinical symptoms manifest. However, the precise neural mechanisms underlying this protective effect, particularly in terms of structural brain changes in individuals with established cognitive impairment, remain an active area of investigation. While some studies in AD cohorts have reported larger brain volumes in certain subcortical structures for bilingual speakers, such as the thalamus, ventral diencephalon, and brainstem,[18] or slower rates of parenchymal volume decline,[19] others have found mixed results. For example, depending on the study, findings of less vs. more gray matter have been reported in areas such as the hippocampal regions.[20,21] This suggests a complex relationship that may depend on the specific brain regions, disease stage, and individual bilingualism characteristics. These mixed findings highlight the need for further investigation into neural differences between monolingual and bilingual speakers with specific neurodegenerative syndromes.

Among the various forms of dementia, primary progressive aphasias (PPAs) offer a unique lens through which to examine the impact of bilingualism on brain structure, given their primary manifestation as a language impairment. Logopenic variant PPA (lvPPA), in particular, is characterized by word-finding difficulties, impaired sentence repetition, and phonological errors, with atrophy typically centered in the left posterior temporoparietal junction, including the inferior parietal lobule and posterior superior temporal gyrus.[22] In most cases, the underlying neuropathology is Alzheimer’s disease,[23] and lvPPA is therefore often considered a language variant of AD.[24] Given the extensive neural networks involved in bilingual language processing, it is plausible that bilingualism might differentially influence the structural integrity of these language-critical regions in lvPPA. While research on structural brain differences in bilingual individuals with lvPPA is still emerging, clinical studies have indicated that bilingual speakers with lvPPA experience a delay in symptom onset compared to monolinguals.[25] Understanding these potential structural adaptations and their clinical implications in lvPPA is crucial, as the disease directly targets the very neural systems that bilingualism relies heavily upon and potentially strengthens.

Despite the accumulating evidence, a comprehensive understanding of how bilingualism influences gray matter volume and cortical thickness in specific neurodegenerative syndromes, particularly across different AD variants, has yet to be established. Many studies have focused on general AD or mild cognitive impairment, with fewer investigations focusing on distinct clinical phenotypes such as amnestic AD, primarily characterized by memory deficits, and lvPPA, primarily characterized by language deficits. Moreover, to our knowledge, this is the first study directly comparing structural brain differences in bilingual versus monolingual individuals with lvPPA. Furthermore, the regional specificity of these structural differences and their relationship to the clinical presentation and progression disease warrant further examination.

This study aims to address these gaps by conducting a thorough investigation into structural brain differences, specifically gray matter volume, in bilingual and monolingual adults diagnosed with either amnestic AD or logopenic variant PPA. By examining these two distinct clinical syndromes, we seek to elucidate whether the structural brain adaptations associated with bilingualism manifest differently depending on the primary cognitive domain affected by neurodegeneration, thereby contributing to a more refined understanding of reserve, resilience, and resistance and their role in mitigating the impact of neurodegenerative pathology.

## METHODS

### Participants

One hundred thirty-six individuals with a diagnosis of amnestic AD and 88 individuals with logopenic variant PPA were included in this retrospective study. Data were previously collected as part of longitudinal research studies (an NIH Program Project Grant and Alzheimer’s Disease Research Center (ADRC) projects) conducted at the University of California, San Francisco (UCSF). Study procedures were approved by the UCSF Institutional Review Board, and patients consented to the use of their behavioral and MRI data for research purposes. All patients were evaluated by a behavioral neurologist, and diagnosis was reached by a multidisciplinary team applying current diagnostic criteria.[22,26] The current sample was restricted to individuals meeting diagnostic criteria for amnestic AD or lvPPA who had a T1-weighted magnetic resonance image (MRI) of adequate quality. Demographic and other participant characteristics for each speaker group (monolingual and bilingual) are presented in Table 1.

**Table 1.** Participant demographics and neuropsychological assessment data. Significant differences (after FDR-correction) are highlighted in BOLD.

|  | Amnesic AD |  |  |  | lvPPA |  |  |  |
| --- | --- | --- | --- | --- | --- | --- | --- | --- |
|  | All, N | Monolingual (N= 119) | Bilingual (N= 17) | P-value | All, N | Monolingual (N= 67) | Bilingual (N= 21) | P-value |
| Age at evaluation, mean (SD) | 136 | 63.90 (9.60) | 65.30 (9.50) | 0.59 | 88 | **64.30 (8.00)** | **69.50 (9.20)** | **0.03** |
| Sex, N | 136 | 119 | 17 | 0.62 | 88 | 67 | 21 | 0.33 |
| Male | 64 | 55 | 9 |  | 41 | 33 | 8 |  |
| Female | 72 | 64 | 8 |  | 47 | 34 | 13 |  |
| Education, mean (SD) | 135 | 15.50 (3.00) | 16.90 (2.80) | 0.07 | 86 | 16.30 (2.50) | 17.50 (2.20) | 0.05 |
| Race, N | 136 | 119 | 17 | 0.07 | 88 | **67** | **21** | **0.008** |
| White | 113 | 101 | 12 |  | 76 | 62 | 14 |  |
| Asian | 9 | 6 | 3 |  | 1 | 0 | 1 |  |
| Black/African American | 4 | 3 | 1 |  | 2 | 1 | 1 |  |
| Native American | 0 | 0 | 0 |  | 0 | 0 | 0 |  |
| Mixed | 0 | 0 | 0 |  | 2 | 1 | 1 |  |
| Other | 4 | 3 | 1 |  | 1 | 0 | 1 |  |
| Unknown | 6 | 6 | 0 |  | 6 | 3 | 3 |  |
| **General** |  |  |  |  |  |  |  |  |
| MMSE (30) | 125 | 22.10 (5.50) | 22.10 (6.50) | 0.29 | 78 | 19.80 (7.20) | 21.80 (6.70) | 0.74 |
| GDS (30) | 105 | 7.00 (5.20) | 6.50 (6.30) | 0.91 | 65 | 7.20 (4.80) | 6.60 (3.10) | 0.94 |
| CDR Box | 135 | 4.60 (2.20) | 5.00 (2.40) | 0.14 | 87 | 3.60 (2.40) | 3.10 (1.50) | 0.77 |
| **Memory** |  |  |  |  |  |  |  |  |
| CVLT 10 min (9) | 117 | 1.50 (2.10) | 1.20 (1.80) | 0.41 | 69 | 2.60 (2.70) | 2.70 (2.70) | 0.79 |
| Rey recall (17) | 120 | 3.50 (3.80) | 2.00 (1.60) | 0.07 | 76 | 6.00 (4.10) | 6.80 (3.70) | 0.66 |
| **Language** |  |  |  |  |  |  |  |  |
| Sentence repetition (5) | 102 | 4.00 (1.30) | 3.90 (1.50) | 0.34 | 68 | 2.00 (1.30) | 2.50 (1.20) | 0.65 |
| Verbal agility (6) | 99 | 4.90 (1.40) | 5.00 (1.00) | 1.00 | 66 | 3.50 (2.10) | 4.70 (1.60) | 0.22 |
| WRAT-4 | 44 | 57.80 (12.10) | 64.00 (4.30) | 0.81 | 43 | 51.70 (9.20) | 54.80 (11.70) | 0.81 |
| Irregular word reading (6) | 93 | 5.80 (0.60) | 5.80 (0.40) | 0.68 | 64 | 5.00 (1.00) | 5.10 (1.60) | 0.54 |
| Sentence comprehension (5) | 96 | 3.70 (1.50) | 3.80 (1.80) | 0.59 | 65 | 3.20 (1.30) | 3.80 (1.10) | 0.57 |
| PPVT (16) | 104 | 14.10 (1.90) | 14.10 (2.60) | 0.59 | 71 | 13.50 (2.80) | 13.00 (3.30) | 0.23 |
| Animal fluency | 118 | 12.20 (6.10) | 11.40 (6.20) | 0.21 | 74 | 9.50 (6.10) | 9.30 (6.40) | 0.67 |
| BNT (15) | 114 | 12.10 (3.10) | 12.10 (3.40) | 0.68 | 72 | 9.10 (4.20) | 9.40 (4.20) | 0.69 |
| **Visuospatial** |  |  |  |  |  |  |  |  |
| Rey copy (17) | 123 | 11.60 (5.00) | 12.40 (4.90) | 0.86 | 77 | 12.40 (4.70) | 14.70 (2.10) | 0.16 |
| Calculations (5) | 116 | 3.20 (1.50) | 3.50 (1.10) | 0.82 | 73 | 3.20 (1.30) | 3.30 (1.00) | 0.73 |
| VOSP (10) | 103 | 6.80 (2.70) | 8.50 (2.30) | 0.17 | 63 | 7.70 (2.30) | 7.80 (2.50) | 0.24 |
| **Frontal executive** |  |  |  |  |  |  |  |  |
| Trails (seconds) | 90 | 78.00 (38.00) | 85.10 (39.90) | 0.20 | 56 | 79.40 (36.30) | 73.20 (38.90) | 0.77 |
| Trails (lines) | 90 | 10.20 (5.50) | 9.80 (5.90) | 0.30 | 56 | 10.40 (5.30) | 12.30 (3.70) | 0.70 |
| Design fluency | 105 | 5.80 (3.80) | 5.80 (2.80) | 0.48 | 68 | 6.40 (3.40) | 6.50 (3.00) | 0.72 |
| Digits forward | 78 | 3.30 (1.40) | 3.60 (1.70) | 0.78 | 63 | 4.50 (1.20) | 4.90 (1.70) | 0.95 |
| Digits backward | 117 | 5.40 (1.30) | 5.50 (1.40) | 0.89 | 72 | 3.10 (1.00) | 3.50 (1.20) | 0.48 |
| D words | 118 | 9.70 (5.40) | 10.40 (4.80) | 0.49 | 74 | 7.60 (4.60) | 8.80 (5.40) | 0.86 |
| Abstraction (6) | 88 | 2.60 (1.70) | 3.90 (2.00) | 0.12 | 39 | 1.90 (1.60) | 2.80 (1.30) | 0.54 |
| Stroop color naming | 86 | 54.10 (23.90) | 58.70 (23.90) | 0.90 | 55 | 44.20 (18.70) | 49.10 (20.30) | 0.93 |

BNT = Boston Naming Test, CVLT = California Verbal Learning Test, MMSE = Mini-Mental State Examination, PPVT = Peabody Picture Vocabulary Test, VOSP = Visual Object and Space Perception battery. Bold text indicates significant between-group differences. FDR correction applied to each family of assessments.

### Language Groups

Of the 224 individuals included in this study, 38 (17 with AD; 21 with lvPPA) were bilingual speakers. Bilingualism status was determined by two study authors via retrospective chart review, as described in prior studies from our group.[25,27] To summarize, patients were classified as bilingual if their chart indicated that they regularly used two or more languages and/or they had the ability to communicate with native speakers in two languages.[28] A description of the languages spoken in the two cohorts is reported in Supplemental Table A. Conversely, individuals were classified as monolingual if their charts did not state information regarding exposure to or experience with a second language. Participants were excluded if it was not clear whether they were monolingual or bilingual according to the above criteria. More specifically, participants were excluded if their chart revealed that a) they had enrolled in classes to learn a second language for only a few years and did not report any other exposure or ongoing experience with the language outside of the classroom; b) they immigrated to a country that had a different majority language from the country of their previous residence, but it was unclear whether they engaged in formal schooling or were employed in their adopted country in an environment that required usage of the primary language spoken in that country; or c) they reported minimal usage of a second language such that it was unclear if they had attained high proficiency or regularly used two or more languages.

### Neuropsychological Assessment

Participants completed a neuropsychological battery consisting of measures that assessed general cognition, verbal and visual episodic memory, speech and language abilities, visuospatial processing, and frontal/executive functioning.[29] Tasks from the neuropsychological battery were grouped by cognitive domain (i.e., general, memory, language, visuospatial and executive/frontal), as shown in Table 1.

### MRI acquisition and preprocessing

Structural T1-weighted magnetic resonance images were acquired on a 1.5 Tesla, 3 Tesla, or 4 Tesla system. MR images were bias-corrected, segmented into gray matter (GM), white matter (WM) and cerebrospinal fluid (CSF)[30] and normalized to Montreal Neurological Institute (MNI) using an optimized geodesic shooting procedure[31] in CAT12 toolbox via Statistical Parametric Mapping (SPM[32] running in MATLAB version 2022b (*The MathWorks Inc*., *2022*). Gray matter images were modulated with voxel values multiplied by Jacobian determinants resulting from the spatial normalization to preserve relative GM volumes from the original images. Further details on the acquisition and harmonization of the MRI data are reported in the Supplementary Materials.

Regions of interest (ROIs) were defined using the human Brainnetome atlas (BNA)[34] which consists of 210 cortical and 36 subcortical regions bilaterally. Compared to traditional anatomical parcellations (e.g., Desikan atlas), the BNA provides a more neurobiologically informed framework by capturing the fine-grained organization of the brain’s network architecture. It has been widely used in neuroimaging research for ROI-based analysis, connectomics, and network-level comparisons in both healthy and clinical populations. Gray matter volumes were extracted from the participants’ segmented and modulated structural images.

### Statistical Analyses

All analyses were conducted in R version 4.5.1.[35] Demographic data were compared between speaker groups of each diagnostic group using Welch’s t-tests and Fisher’s exact tests. Neuropsychological data were analyzed through multiple linear regression analyses (*stats* package in R) to compare group differences while covarying for sociodemographic factors, namely age, sex, and education.[36] A false discovery rate (FDR) correction was applied to analyses conducted within each cognitive domain. Where violations to normality were observed (QQ plots and Shapiro-Wilk tests), we additionally fitted robust linear regression models (*robustbase* package in R).

Gray matter volumes were analyzed using a series of linear mixed-effects regression models with the dependent variable of gray matter volume and a two-way interaction term of speaker status (mono- and bi-lingual) and ROI, with MMSE and sex as covariates, and a random intercept of participant, within each clinical group. Analyses were conducted in the *lme4* package[37] in R Studio. *P*-values were derived via the *emmeans* package,[38] with an FDR correction applied to ROIs within each hemisphere. To examine whether education contributed to differences in gray matter volume, we conducted exploratory model comparisons by sequentially adding education and the interaction between education and speaker status to the base model, which included speaker status × region of interest (ROI), MMSE and sex as covariates, and a random intercept of participant. Model comparisons were performed separately within each clinical group and hemisphere.

## RESULTS

### Sociodemographic Data and Performance on Neuropsychological Battery

#### Amnestic AD

The AD cohort overall was 47% male with a mean level of education of 16.5 years and 83% of participants self-identifying as White; these demographic variables did not differ by monolingual or bilingual speaker status. The average age at evaluation was 64.5, and the monolingual and bilingual groups also did not differ in terms of disease severity as measured by the MMSE or CDR sum of boxes scores.

On neuropsychological testing (Table 1), no statistically significant differences were observed between monolingual and bilingual participants on any of the neuropsychological tests, after controlling for age, sex, and education.

#### Logopenic variant PPA

The lvPPA cohort was 47% male with 17 mean years of education, which did not differ between monolingual and bilingual groups. The speaker groups also had similar MMSE and CDR sum of boxes scores. The monolingual and bilingual speakers did differ with respect to race, with 76% of the monolingual speakers and 24% of the bilingual speakers self-identifying as White (*p* = 0.008), and the bilingual speakers were older at evaluation (monolinguals mean = 64 years; bilinguals mean = 70 years; *p* = 0.03).

On neuropsychological testing, no statistically significant differences were observed between monolingual and bilingual participants on any of the neuropsychological tests, after controlling for age, sex, and education.

### Volumetric Results

#### Amnestic AD

Results revealed a distinct pattern of reduced gray matter volume in bilingual individuals with amnestic AD compared to monolinguals (Table 2; Figure 1). These differences were most prominent in the fusiform gyrus, where bilinguals showed less volume across multiple subregions bilaterally. Additional differences (less volume in bilinguals) were found in the hippocampus, particularly in the rostral and caudal portions, as well as in areas of the lateral occipital cortex, including the middle occipital gyrus, occipital pole, and superior occipital gyrus. In the right hemisphere, bilinguals also exhibited less volume in the inferior parietal lobule, middle temporal gyrus, and several subregions of the medial ventral occipital cortex, such as the lingual gyrus, cuneus, and parieto-occipital sulcus. Finally, less volume in bilinguals was observed in the precuneus and the caudal portion of the superior temporal gyrus. These findings suggest a widespread pattern of volumetric loss across temporal, parietal, and occipital regions.

**Table 2.** Regions showing significant (FDR-corrected) decrease in gray matter volume in bilinguals compared to monolinguals with Amnestic AD.

| Hemisphere | ROI | estimate | SE | z.ratio | p.value |
| --- | --- | --- | --- | --- | --- |
| left | Fusiform gyrus - rostroventral | 0.343 | 0.092 | 3.745 | 0.000 |
| left | Fusiform gyrus - medioventral | 0.361 | 0.092 | 3.939 | 0.000 |
| left | Fusiform gyrus - lateroventral | 0.256 | 0.092 | 2.797 | 0.005 |
| left | Hippocampus - rostral | 0.200 | 0.092 | 2.182 | 0.029 |
| left | Lateral occipital cortex - middle occipital gyrus | 0.183 | 0.092 | 1.996 | 0.046 |
| left | Lateral occipital cortex - superior occipital gyrus | 0.244 | 0.092 | 2.663 | 0.008 |
| right | Fusiform gyrus - rostroventral | 0.374 | 0.098 | 3.805 | 0.000 |
| right | Fusiform gyrus - medioventral | 0.347 | 0.098 | 3.539 | 0.000 |
| right | Fusiform gyrus - lateroventral | 0.309 | 0.098 | 3.143 | 0.002 |
| right | Hippocampus - rostral | 0.213 | 0.098 | 2.165 | 0.030 |
| right | Hippocampus - caudal | 0.207 | 0.098 | 2.109 | 0.035 |
| right | Inferior parietal lobule - area 39 | 0.201 | 0.098 | 2.049 | 0.040 |
| right | Inferior parietal lobule - area 40 | 0.223 | 0.098 | 2.272 | 0.023 |
| right | Lateral occipital cortex - middle occipital gyrus | 0.242 | 0.080 | 3.041 | 0.002 |
| right | Lateral occipital cortex - occipital pole | 0.236 | 0.080 | 2.965 | 0.003 |
| right | Lateral occipital cortex - middle occipital gyrus | 0.235 | 0.098 | 2.394 | 0.017 |
| right | Lateral occipital cortex - superior occipital gyrus | 0.418 | 0.098 | 4.262 | 0.000 |
| right | Middle temporal gyrus - dorsolateral | 0.203 | 0.098 | 2.067 | 0.039 |
| right | Middle temporal gyrus - ventrolateral | 0.207 | 0.098 | 2.104 | 0.035 |
| right | Medial ventral occipital cortex - parieto-occipital sulcus | 0.249 | 0.098 | 2.537 | 0.011 |
| right | Medial ventral occipital cortex - occipital gyrus | 0.289 | 0.098 | 2.945 | 0.003 |
| right | Medial ventral occipital cortex - cuneus | 0.227 | 0.098 | 2.313 | 0.021 |
| right | Medial ventral occipital cortex - lingual gyrus | 0.361 | 0.098 | 3.678 | 0.000 |
| right | Medial ventral occipital cortex - cuneus | 0.247 | 0.098 | 2.519 | 0.012 |
| right | Precuneus | 0.318 | 0.098 | 3.240 | 0.001 |
| right | Superior temporal gyrus - caudal | 0.209 | 0.098 | 2.127 | 0.033 |

**Figure 1.**
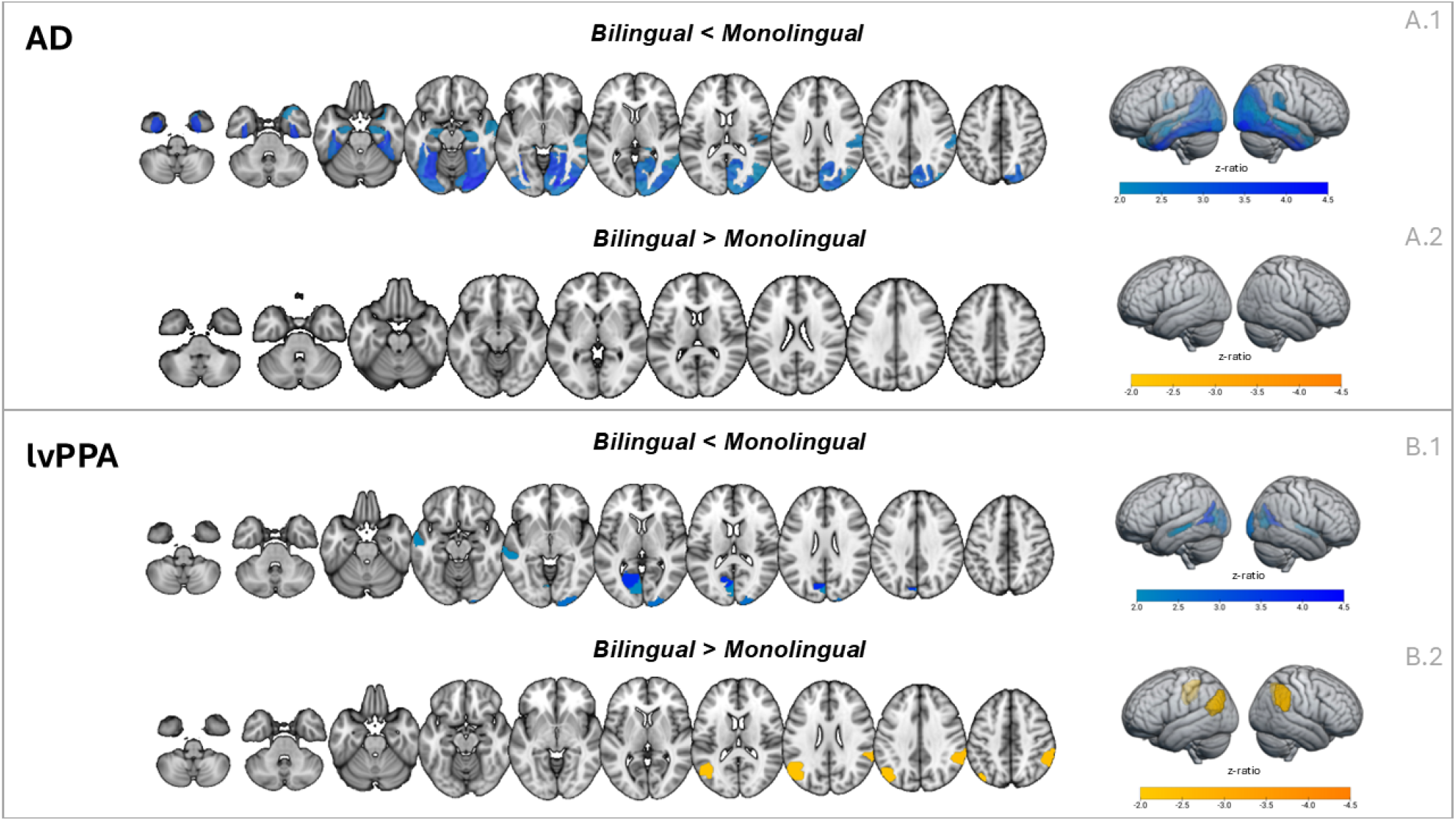
Significant (FDR-corrected) volumetric differences between bilingual and monolingual individuals in the two clinical groups (lvPPA, AD). The maps show z ratio values resulting from the linear mixed-effect model analyses (reported in Table 1-4). Regions showing less GMV in bilinguals than in monolinguals are shown in blue, while regions showing greater GMV in bilinguals are shown in orange. Images were created with MRIcroGL.

#### Logopenic variant PPA

Bilingual individuals with lvPPA showed less volume than monolinguals in several left hemisphere regions, including the ventrolateral middle temporal gyrus, the ventromedial occipital gyrus, the cuneus, and the caudal portion of the superior temporal gyrus (Table 3). In the right hemisphere, bilinguals demonstrated less volume in the middle occipital gyrus and again in the caudal superior temporal gyrus. These differences suggest that bilinguals with lvPPA show a pattern of volumetric loss in posterior temporal and occipital regions, as well as in parts of the visual association cortex.

**Table 3.**
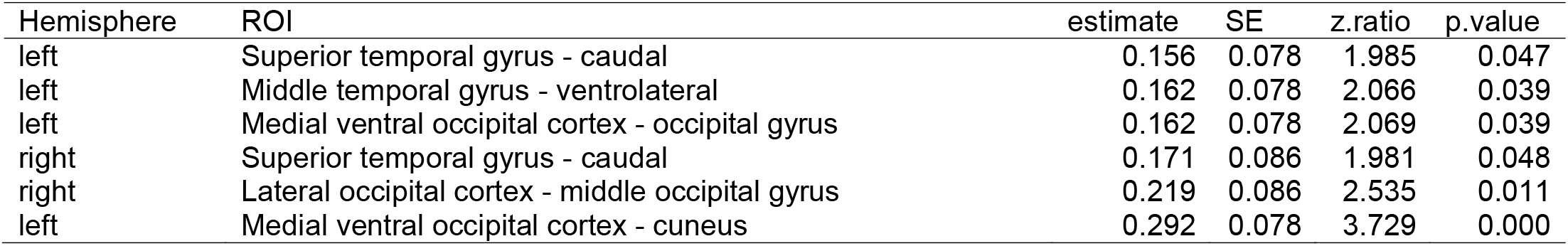
Regions showing significant (FDR-corrected) decrease in gray matter volume in bilinguals compared to monolinguals with lvPPA.

Bilingual individuals with lvPPA showed greater volume in specific regions of the inferior parietal lobule in both hemispheres (Table 4). These included the caudoventral subregion on the left and the rostroventral subregion on the right. These parietal areas are often associated with attention and language integration, inclusive of phonology.

**Table 4.** Regions showing significant (FDR-corrected) increase in gray matter volume in bilinguals compared to monolinguals with lvPPA.

| Hemisphere | ROI | estimate | SE | z.ratio | p.value |
| --- | --- | --- | --- | --- | --- |
| right | Inferior parietal lobule - rostroventral | -0.210 | 0.086 | -2.428 | 0.015 |
| left | Inferior parietal lobule - caudoventral | -0.179 | 0.078 | -2.277 | 0.023 |

### Exploratory Analysis: Effect of Education

In the amnestic AD group, exploratory analyses including education as an additional predictor significantly improved model fit for the left hemisphere data (χ^2^(1) = 5.04, *p* =.025). Adding the education × speaker status interaction did not yield further improvement (χ^2^(1) = 2.36, *p* =.124). No significant improvement was observed for right hemisphere data in this group.

For individuals with lvPPA, model comparisons showed no evidence that including education or its interaction with speaker status improved model fit in either hemisphere (*ps* >.16). These results indicate that, among the covariates examined, education may have a different impact in the two clinical variants, by modestly explaining variance in left hemisphere volume in the amnestic AD group, but not in lvPPA.

## DISCUSSION

In this study, we observed differences in brain volume between monolingual and bilingual speakers with two clinical variants of AD. Despite similar scores on neuropsychological testing, bilingual speakers with amnestic AD had less volume in certain brain regions compared to monolingual speakers, including the right IPL, right STG, right MTG, right precuneus, and bilateral hippocampi, fusiform gyrus, and occipital cortex. Correspondingly, bilingual and monolingual individuals with lvPPA performed similarly on neuropsychological measures, yet bilinguals with lvPPA had less volume in the bilateral STG, left MTG, and bilateral occipital regions. Furthermore, the bilingual group with lvPPA attained similar cognitive scores despite also being significantly older. Since bilingual individuals in both AD variants acheived comparable cognitive outcomes with less structural brain support, these results are in line with previous models of cognitive reserve.[2–4,6] These findings support the theory that bilingualism may help maintain cognitive functioning despite age-related or disease-related brain changes, thus promoting cognitive resilience.

Our results generally are in line with previous studies showing reduced brain volumes in bilingual speakers in regions associated with AD, including parahippocampal[21] and mesial temporal regions.[39,40] Despite these reductions, bilingual individuals can maintain cognitive function[21] and perform better on cognitive measures when the degree of brain atrophy is similar to monolingual speakers.[41] However, previous studies have varied in their findings, with some showing greater volume in certain regions for bilingual speakers with AD, including orbitofrontal,[42] hippocampal,[20] thalamic, ventral diencephalon, and brainstem regions.[18] Therefore, there remains a need for further, larger studies on bilingualism and AD that can help clarify these mixed findings.

Interestingly, bilingual speakers with lvPPA additionally had larger volumes in bilateral IPL compared to monolingual speakers. Similar instances of regionally-specific brain reserve in bilateral IPL have also been reported in healthy older bilingual speakers,[43,44] demonstrating that this regional difference tends to persist at older ages in comparison to frontal regions that are also initially larger in bilingual speakers.^[44,45]^ Therefore, our finding of higher volume in bilateral IPL in lvPPA bilingual speakers may reflect that these regions continue to exhibit brain reserve longer than other regions that may initially also be larger in healthy bilingual speakers. The higher volume in the bilateral IPL is also interesting to consider in light of previous findings of an older age of symptom onset in lvPPA but not in amnestic AD. Since the left IPL is often considered the epicenter of disease in lvPPA,[46–48] the neuroprotective effects of bilingualism in the IPL may promote resistance in AD, by helping bolster against the initiation and spread of neurodegenerative changes associated with lvPPA that begin in this region, resulting in a later age at symptom onset.[25]

In summary, our findings demonstrate that bilingualism may play a role in cognitive reserve in both amnestic AD and lvPPA, with additional contributions to brain reserve in lvPPA, specifically in the bilateral IPL. Our findings may therefore be an important step toward understanding which effects of bilingualism are domain-general versus domain-specific within cognitive domains and clinical syndromes. Findings may also help address previously mixed findings in AD in terms of its protective effect in cognitive reserve and resilience by clarifying whether speaking two or more languages has differential effects that depend on associations with distinct AD variants.

Strengths of our study included the comprehensive phenotyping of our participants in terms of symptoms and neuropsychological testing, which allowed for detailed study of monolingual and bilingual speakers with two different clinical AD variants. In addition, there are few studies that have previously evaluated neuroimaging in bilingual participants across AD variants.

There are several important limitations to consider when interpreting the results from this study. It is important to recognize that findings regarding brain volume depend on bilingualism factors, including L2 age at acquisition, proficiency, exposure, and linguistic distance of the languages,[9,43,49] and these likely also lead to dynamic changes in a bilingual brain, which cannot be fully captured in a cross-sectional study.[14] Therefore, future studies should consider these factors in interpreting results. Furthermore, there is a need for future studies that examine longitudinal data, which are essential to provide robust evidence for cognitive and brain reserve. Nevertheless, we hope that these results can serve as a starting point for future longitudinal studies that account for these important bilingualism factors.

## Supporting information

MRI data acquisition and harmonization; Supplemental Table A

## Data Availability

All data produced in the present study are available upon reasonable request to the authors

## ACKNOWLEDGEMENTS

The authors have no competing interests to declare. This work was supported by the National Institutes of Health under the following grants: R01AG080396, K23DC018021, and P01AG019724 (to JD); the National Institute on Aging, grant R01AG080470 (to SMG); the National Institute on Deafness and Other Communication Disorders, grants R01DC016291 (to MLH) and K24DC015544 (to MLGT); and the National Institute of Neurological Disorders and Stroke, grant R01NS050915 (to MLGT).

## CONSENT STATEMENT

The participants and/or authorized decision-making surrogate provided written consent for this study. Approval for the study was obtained from the UCSF Institutional Review Board for human research.

