## Supplementary material for "Cognitive and brain reserve in bilingual speakers with clinical AD variants": MRI data acquisition and harmonization; Supplemental Table A

Structural T1-weighted magnetic resonance images were acquired on a 1.5 Tesla (Magnetom VISION system (Siemens, Iselin, NJ); slice thickness = 1.5 mm; field of view = 256 mm2; matrix 256 × 256; voxel size 1.0 × 1.5 × 1.0 mm3; TR= 10 ms; TE = 4 ms; flip angle = 15°), 3 Tesla (Trio/Prisma; slice thickness = 1 mm, field of view = 256 mm2, matrix = 240 × 256, voxel size = 1mm3 isotropic, TR = 2300 ms, TE = 2.9 ms, flip angle = 9°), or 4 Tesla system (Bruker MedSpec system with an 8 channel head coil controlled by a SiemensTrio console; slice thickness 1mm; FOV = 256 mm2, matrix = 256 × 256, voxel size = 1mm3, TR = 2300 ms, TE = 3 ms, flip angle = 7°).

To mitigate biases introduced by the use of different scanners and sequence parameters, we applied the ComBat method (Johson et al., 2007) to the GM volumes. This approach utilizes a multivariate linear mixed-effects regression that incorporates terms for both biological variables (age and total intracranial volume) and scanner types to adjust for imaging feature measurements. ComBat harmonization analyses were conducted using a publicly available R package, accessible at <https://github.com/Jfortin1/ComBatHarmonization>.

***Reference:***

Johnson, W. E., Li, C., & Rabinovic, A. (2007). Adjusting batch effects in microarray expression data using empirical Bayes methods. *Biostatistics (Oxford, England)*, *8*(1), 118–127. https://doi.org/10.1093/biostatistics/kxj037

**Supplemental Table A.** L1 and L2 for bilingual speakers by group. Speakers who acquired English as a third or fourth language are indicated with * and **, respectively.

Amnestic AD (N = 17)

| L1 | L2 | N |
| --- | --- | --- |
| Cantonese | English | 1 |
| Chinese | English | 1 |
| English | French | 6 |
| English | German | 1 |
| English | Spanish | 1 |
| English | Swiss German | 1 |
| Hungarian | German | 1** |
| Spanish | English | 3 |
| Tagalog | English | 1 |
| Tigrinya | English | 1 |

lvPPA (N = 21)

| L1 | L2 | N |
| --- | --- | --- |
| Arabic | French | 1* |
| Cantonese | English | 1 |
| English | French | 1 |
| English | German | 2 |
| English | Greek | 1 |
| English | Spanish | 5 |
| German | English | 1 |
| Greek | English | 1 |
| Hebrew | English | 1 |
| Italian | English | 1 |
| Malayalam | Hindi | 1* |
| Marathi | Hindi | 1* |
| Portuguese | English | 1 |
| Portuguese | French | 1* |
| Spanish | English | 1 |
| Turkish | English | 1 |
